# Development of a High-Performance Multiparametric MRI Oropharyngeal Primary Tumor Auto-Segmentation Deep Learning Model and Investigation of Input Channel Effects: Results from a Prospective Imaging Registry

**DOI:** 10.1101/2021.07.27.21261114

**Authors:** Kareem A. Wahid, Sara Ahmed, Renjie He, Lisanne V. van Dijk, Jonas Teuwen, Brigid A. McDonald, Vivian Salama, Abdallah S.R. Mohamed, Travis Salzillo, Cem Dede, Nicolette Taku, Stephen Y. Lai, Clifton D. Fuller, Mohamed A. Naser

**Affiliations:** Department of Radiation Oncology, University of Texas MD Anderson Cancer Center, Houston, TX; Department of Medical Imaging, Radboud University Medical Centre, Nijmegen, The Netherlands; Department of Head and Neck Surgery, University of Texas MD Anderson Cancer Center, Houston, TX

## Abstract

**Background and Purpose:** Oropharyngeal cancer (OPC) primary gross tumor volume (GTVp) segmentation is crucial for radiotherapy. Multiparametric MRI (mpMRI) is increasingly used for OPC adaptive radiotherapy but relies on manual segmentation. Therefore, we constructed mpMRI deep learning (DL) OPC GTVp auto-segmentation models and determined the impact of input channels on segmentation performance.

**Materials and Methods:** GTVp ground truth segmentations were manually generated for 30 OPC patients from a clinical trial. We evaluated five mpMRI input channels (T2, T1, ADC, Ktrans, Ve). 3D Residual U-net models were developed and assessed using leave-one-out cross-validation. A baseline T2 model was compared to mpMRI models (T2+T1, T2+ADC, T2+Ktrans, T2+Ve, all 5 channels [ALL]) primarily using the Dice similarity coefficient (DSC). Sensitivity, positive predictive value, Hausdorff distance (HD), false-negative DSC (FND), false-positive DSC, surface DSC, 95% HD, and mean surface distance were also assessed. For the best model, ground truth and DL-generated segmentations were compared through a Turing test using physician observers.

**Results:** Models yielded mean DSCs from 0.71 (ALL) to 0.73 (T2+T1). Compared to the T2 model, performance was significantly improved for HD, FND, sensitivity, surface DSC, and 95% HD for the T2+T1 model (p<0.05) and for FND for the T2+Ve and ALL models (p<0.05). There were no differences between ground truth and DL-generated segmentations for all observers (p>0.05).

**Conclusion:** DL using mpMRI provides high-quality segmentations of OPC GTVp. Incorporating additional mpMRI channels may increase the performance of certain evaluation metrics. This pilot study is a promising step towards fully automated MR-guided OPC radiotherapy.

## 1. Introduction

Oropharyngeal cancer (OPC), a type of head and neck squamous cell carcinoma (HNSCC), is among the most common malignancies globally [1]. Treatment for OPC often includes radiotherapy because of its high cure rate [2]. Segmentation (also termed contouring) of the primary gross tumor volume (GTVp) on radiologic imaging is necessary for the OPC radiotherapy workflow. The GTVp, with a clinical and planning safety margin, acts as a target volume to deliver the radiotherapy dose. Therefore, inadequate GTVp definition may cause under-dosage of the tumor or over-dosage of surrounding normal tissues [3,4]. However, the current clinical standard is manual segmentation by physician experts, which is labor-intensive and subject to high inter-observer variation [5–7]. Therefore, an auto-segmentation tool would be a promising alternative to the current manual standard in OPC radiotherapy workflows.

Deep learning (DL) has found wide success in auto-segmentation [8,9], with many HNSCC auto-segmentation studies applying DL to CT imaging [10–12]. Although CT is the most commonly used imaging modality in OPC radiotherapy planning, MRI has been increasingly recognized as essential for tumor segmentation because of its exceptional soft-tissue contrast [13,14]. Additionally, the emergence of MR-Linac technology, an image-guided adaptive radiotherapy approach, has further incentivized the incorporation of MRI in OPC radiotherapy planning. Importantly, we recently demonstrated the utility of DL for HNSCC organ-at-risk auto-segmentation using MRI, with improvements in performance, execution time, and dosimetric differences compared to other auto-segmentation methods [15]. While several DL tumor auto-segmentation studies for nasopharyngeal cancer using MRI have been published [16–25], to our knowledge, only one study has been published for OPC [26]. Since HNSCC tumors at different anatomical sites have distinct anatomic boundaries and characteristics [27,28], it is crucial that tumor segmentation models are developed for each site accordingly. Consequently, there exists an unmet need for OPC DL tumor segmentation tools using MRI.

Multiparametric MRI (mpMRI) incorporates multiple sequence acquisitions that highlight anatomical and functional information in tumors. For example, dynamic contrast-enhanced (DCE) MRI and diffusion-weighted imaging (DWI) can quantify tumor perfusion and diffusion patterns, respectively, and may affect OPC treatment guidance [29,30]. Recent studies of PET/CT OPC DL auto-segmentation [11,20,31–35] have demonstrated increased segmentation performance when combining functional and anatomical modalities. However, investigations that combine anatomical with functional MRI in HNSCC to achieve acceptable DL auto-segmentation performance are lacking [36,37].

In this pilot study, we evaluated the effects of anatomical and functional mpMRI inputs on OPC GTVp segmentation performance. Using open-source DL frameworks with standardized clinical trial data, we trained and evaluated DL models based on variable mpMRI input channels. We then compared the models qualitatively and quantitatively to determine which channel combinations led to the best segmentation results. Finally, we characterized the clinical acceptability of the best-performing model using physician experts.

## 2. Methods

### 2.1. Imaging Data

We acquired pre-radiotherapy T2-weighted (T2), contrast-enhanced T1-weighted Dixon fat-suppressed (T1), DCE, and DWI MRI sequences in Digital Imaging and Communications in Medicine (DICOM) format for 124 HNSCC patients from a prospective clinical trial investigating longitudinal mpMRI (NCT03145077). Images were collected from August 2018-August 2019 under a HIPAA-compliant protocol approved by The University of Texas MD Anderson Cancer Center’s IRB (RCR03-0800). The protocol included a waiver of informed consent. We curated 30 OPC patients with a visible GTVp based on the complete availability of T2, T1, DCE, and DWI image sets (**Fig. S1**). Demographic characteristics of the patients are shown in **Table S1**. Imaging was performed on a Siemens Aera scanner with a magnetic field strength of 1.5 T and standardized acquisition parameters (**Table S2**). All patients were immobilized with a thermoplastic mask. Apparent diffusion coefficient (ADC) parametric maps were derived from DWI sequences through a proprietary Siemens algorithm (Munich, Germany) using a monoexponential model. The Tofts model was used to generate parametric maps from DCE sequences for the volume transfer constant (Ktrans) and the extravascular extracellular volume fraction (Ve) [38]. Additional details regarding DCE parametric map generation can be found in our previous publication [39]. GTVp structures were manually segmented in the DICOM-RT Structure format by a physician (radiologist with >5 years of expertise in HNSCC) in Velocity AI v.3.0.1 (Atlanta, GA, USA). GTVp structures were segmented on the T2 MRI, but the physician could consult the other images. An example of the mpMRI images used in this study and overlying GTVp segmentation for one patient is shown in **Figure 1A**.

**Figure 1.**
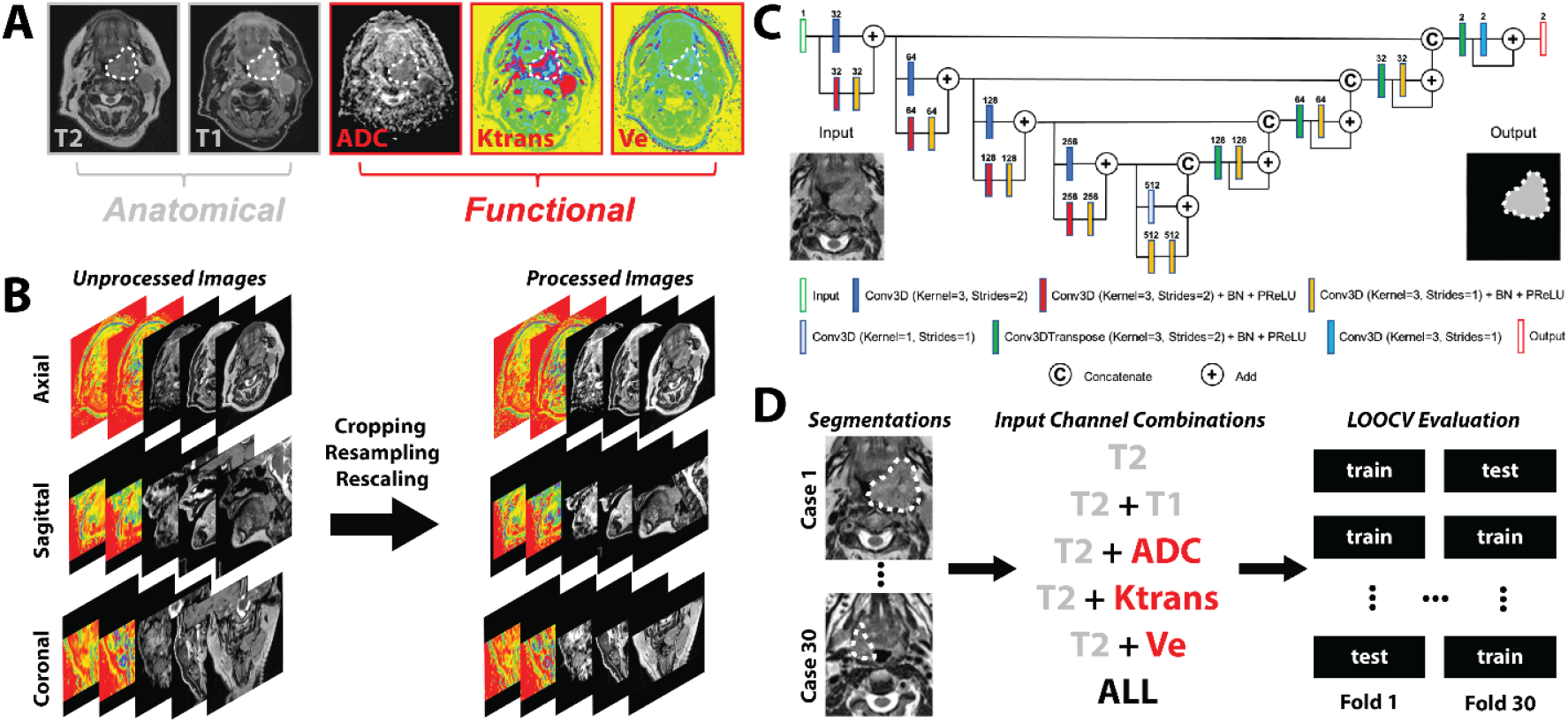
Annotation, processing, and analysis of data used in this study. **(A)** Multiparametric MRI input channels for oropharyngeal tumor segmentation. The white dotted line depicts the primary gross tumor volume segmentation. Anatomical sequence images are outlined in grey boxes, while functional sequence parametric map images are outlined in red boxes. **(B)** Image processing steps which included image cropping, resampling, and rescaling. **(C)** An illustration of the 3D Residual U-net model architecture. For illustrative purposes, only one input channel (T2-weighted image) is shown, but multiple input channel combinations were used throughout the analysis as separate models. **(D)** Overall study design which incorporated multi-channel input combinations coupled to a leave-one-out cross-validation (LOOCV) evaluation approach. T2=T2-weighted MRI, T1=T1-weighted MRI, ADC=apparent diffusion coefficient, Ktrans=volume transfer constant, Ve=extravascular extracellular volume fraction, ALL=all 5 input channels. BN=Batch normalization, PReLU=parametric rectified linear unit activation function.

### 2.2. Image Processing

To ensure adequate MRI comparability between patients [40], we performed intensity standardization for all images. Anatomical sequences (T2, T1) were standardized using a Z-score (mean=0, standard deviation=1), while functional parametric maps (ADC, Ktrans, Ve) were truncated to the 10th and 90th percentile for all patients and rescaled to [-1, 1] as per a previous study [36]. All images were cropped to the smallest field of view (Ktrans, Ve) and resampled to the T2 resolution. An example of the image processing workflow is shown in **Figure 1B**.

### 2.3. Segmentation Model Architecture and Implementation

A DL convolutional neural network based on the 3D Residual U-net architecture [41,42] was implemented in the Medical Open Network for Artificial Intelligence (MONAI) software package [43] (**Fig. 1C**). The GTVp mask was used as the ground truth target to train the segmentation model. The MRI images acted as variable-channel inputs to the models. We investigated the following channel combinations as separate models: T2, T2+T1, T2+ADC, T2+Ktrans, T2+Ve, and all five input channels (ALL). The T2 model acted as a baseline of comparison for all other models. We implemented an Adam optimizer with a Sørensen-Dice similarity coefficient (DSC) loss function. The models were trained for 700 iterations with a learning rate of 2 × 10^−4^ for the first 550 iterations and 1 × 10^−4^ for the remaining 150 iterations. Data augmentation was used to mitigate overfitting. Additional details on the DL architecture and implementation are found in **Supplementary Methods**.

### 2.4. Model Evaluation

Model performance was primarily assessed using DSC. We also implemented additional spatial similarity metrics, including Hausdorff distance (HD), false-negative DSC (FND), false-positive DSC (FPD), sensitivity, positive predictive value (PPV), surface DSC, 95% HD, and mean surface distance (MSD). For surface DSC, a tolerance of 3.0 mm was selected as suitable from previous inter-observer variability studies on T2 MRI of OPC GTVp [44]. Surface distance metrics were calculated using the surface-distance Python package [45], while all other metrics were calculated in Elekta ADMIRE v.2.9 (Stockholm, Sweden). Each model was trained and evaluated using leave-one-out cross-validation (LOOCV) (**Fig. 1D**).

### 2.5. Clinical Evaluation

For our best-performing model, we assigned three physician expert observers (radiologist from *2.1* >1-year post-segmenting, two radiation oncologists) to evaluate the ground truth and corresponding DL-generated segmentations using subjective scoring criteria based on a 4-point Likert scale. The score categories were: 1 = requires corrections, large errors; 2 = requires corrections, minor errors; 3 = clinically acceptable, errors not clinically significant; 4 = clinically acceptable, highly accurate. Additionally, we asked observers to predict the source of the segmentations as either human (ground truth) or DL-generated through a modified Turing test [46]. Ground truth and DL-generated segmentations for all 30 patients were anonymized and randomly presented to experts for clinical evaluation. Experts were blinded to the segmentation source.

### 2.6. Statistical Analysis

After performing a Shapiro-Wilk test, we found that our data were not normally distributed (p<0.05); therefore, we utilized nonparametric statistical tests. We used one-sided Wilcoxon signed-rank tests (alternative hypothesis of greater than for DSC, sensitivity, surface DSC, and PPV; alternative hypothesis of less than for HD, FND, FPD, 95% HD, and MSD) to evaluate differences between our baseline T2 model and models with additional channels. We used Mann-Whitney U tests to detect differences in model performance based on tumor subsite (base of tongue vs. tonsil). Additionally, to assess correlations of tumor size with model performance, we calculated Pearson correlation coefficients with corresponding p-values of ground truth volume against DSC, HD, and surface DSC for every model. Finally, to assess the clinical evaluation of ground truth against DL-generated segmentations, for each observer we implemented a two-sided Wilcoxon signed-rank test for scores and a McNemar test for source predictions. For all statistical analyses, p-values less than 0.05 were considered significant. Analyses were performed in Python v.3.7.9. Code notebooks can be found at GitHub (https://github.com/kwahid/mpMRI_OPC_GTVp_segmentation).

## 3. Results

### 3.1. Model Performance

**Figure 2** shows boxplots of model performance for all tested input channel combinations with respect to different evaluation metrics. T2+T1 was the best performing model overall with the best mean scores in DSC, HD, sensitivity, surface DSC, and 95% HD. ALL was the worst performing model overall with the worst mean scores in in DSC, FPD, and PPV. T2 performed best in FPD and PPV, but worst in sensitivity and 95% HD. T2+Ve performed best in FND and MSD but worst in HD. T2+Ktrans performed worst in MSD. T2+T1 had significantly better performance (p<0.05) than the baseline T2 model for HD, FND, sensitivity, surface DSC, and 95% HD. T2+Ve and ALL had significantly better performance (p<0.05) than the baseline T2 model for FND. **Figure S2** shows a heatmap of p-values comparing channel combinations to the baseline T2 model. A subgroup analysis revealed no significant differences in model performance for any combination of models and metrics based on OPC subsite, as all p-values were > 0.05 (**Table S3**).

**Figure 2.**
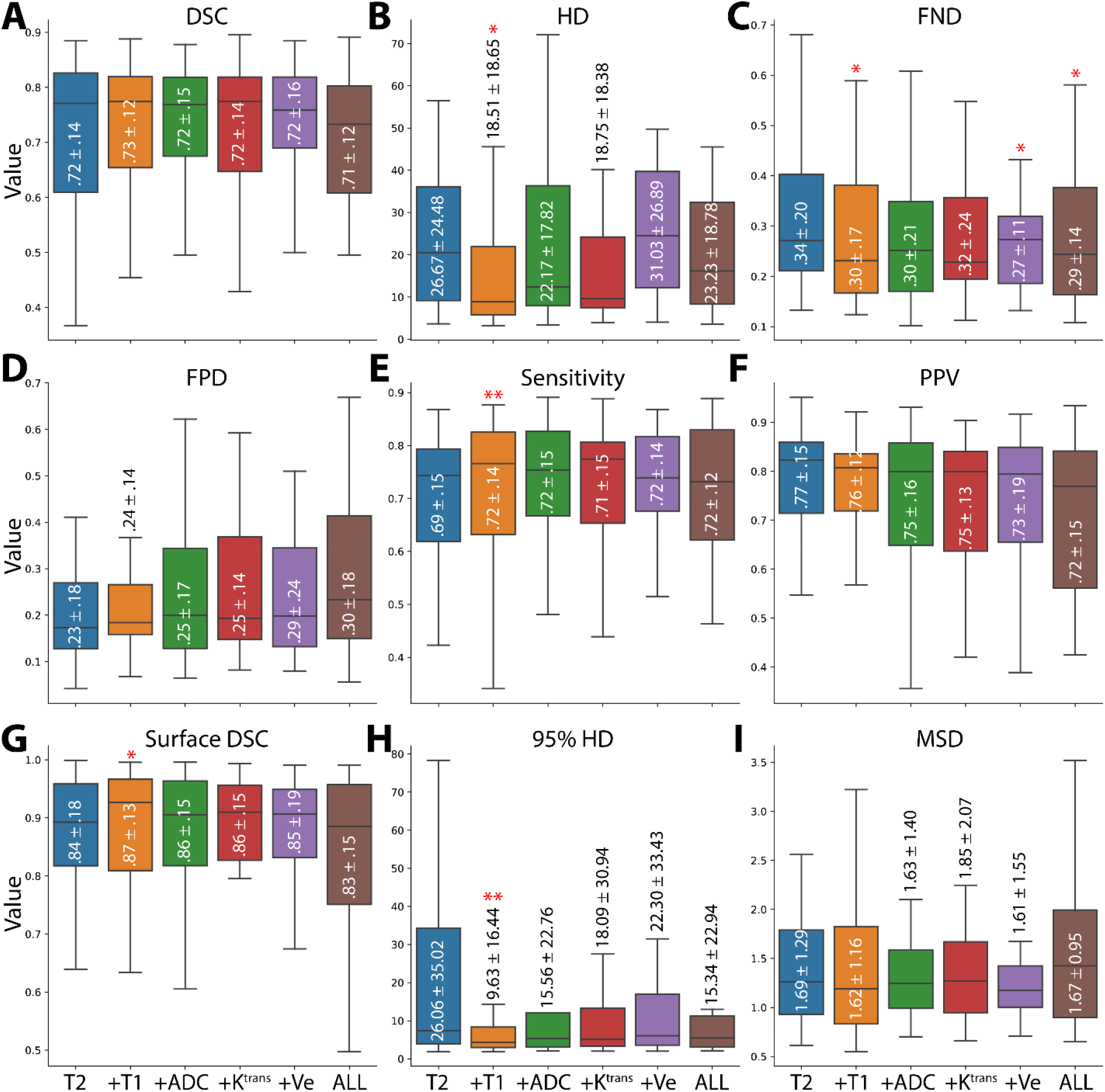
Boxplots comparing evaluation metrics of models built with different input channels. Evaluation metrics correspond to Dice similarity coefficient (DSC) (**A**), Hausdorff distance (HD) (**B**), false-negative DSC (FND) (**C**), false-positive DSC (FPD) (**D**), sensitivity (**E**), positive predictive value (PPV) (**F**), surface DSC (**G**), 95% HD (**H**), and mean surface distance (MSD) (**I**). Boxes show quartiles and median lines, while whiskers extend to the remaining distribution. Mean ± standard deviation is shown inside or adjacent to the corresponding box. The single and double stars above the boxplots correspond to significantly lower or higher values, respectively, compared to the baseline model for that metric. T2=T2-weighted MRI, T1=T1-weighted MRI, ADC=apparent diffusion coefficient, Ktrans=volume transfer constant, Ve= extravascular extracellular volume fraction, ALL=all 5 input channels.

**Figure 3** shows examples of model segmentations compared to ground truth segmentations for high-, medium-, and low-performance cases, based on DSC scores across all models. For the high-performance case, the T2 model demonstrates a DSC of 0.88, with the incorporation of additional channels leading to DSC scores of 0.87-0.90. For the medium-performance case, the T2 model demonstrates a DSC of 0.71, with the incorporation of additional channels leading to DSC scores of 0.72-0.78. For the low-performance case, the T2 model demonstrated a DSC of 0.37 and many spuriously predicted voxels in the posterior region of the head, with the incorporation of additional channels reducing the number of spurious voxels and leading to DSC scores of 0.52-0.61.

**Figure 3.**
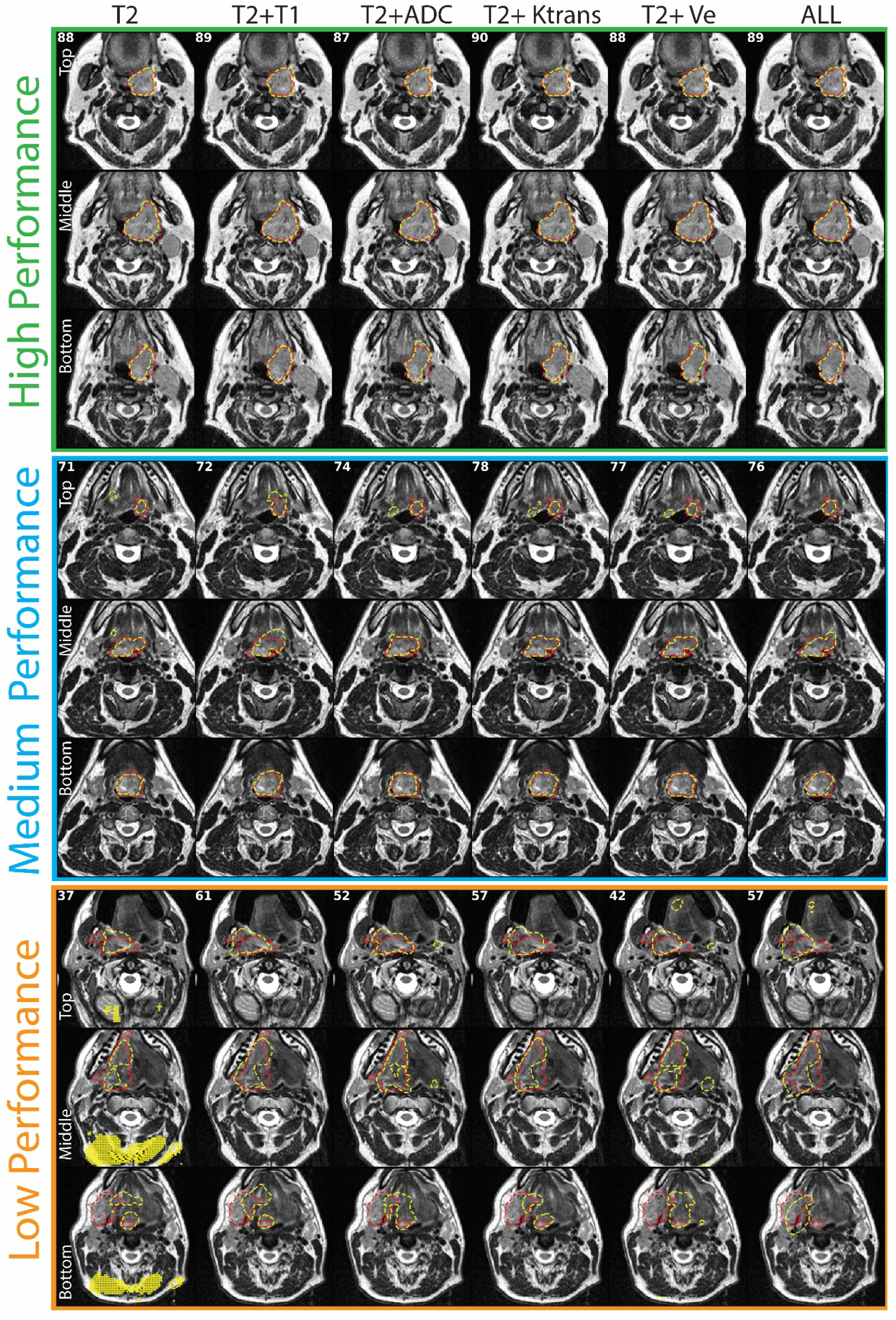
2D axial slice representations of ground truth segmentations (red dotted outline) and predicted segmentations (yellow dotted outline) for high- (green), medium- (blue), and low- (orange) performance cases. Slices for each case are shown in rows superiorly to inferiorly (top, middle, and bottom). Models are shown in columns. The DSC scores for corresponding models are shown in the top left corners. The high-performance case corresponds to a left tonsillar T4 tumor. The medium-performance case corresponds to a left base of tongue T4 tumor. The low-performance case corresponds to a right base of tongue T4 tumor. T2=T2-weighted MRI, T1=T1-weighted MRI, ADC=apparent diffusion coefficient, Ktrans=volume transfer constant, Ve=extravascular extracellular volume fraction, ALL=all 5 input channels.

### 3.2. Size Dependence of Models

**Figure 4** shows correlation graphs of the various models comparing tumor size to DSC, HD, and surface DSC. The range of values for tumor size were 1.74-45.19cc. Every model showed non-significant positive correlations for DSC (p>0.05) and significant positive correlations for HD (p<0.005), except for T2+Ve (r=0.33, p=0.079) and ALL (r=0.06, p=0.76). Every model also showed significant negative correlations for surface DSC (p<0.05), except for T2+ADC (r=-0.34, p=0.07), T2+Ve (r=-0.30, p=0.11), and ALL (r=-0.30, p=0.1).

**Figure 4.**
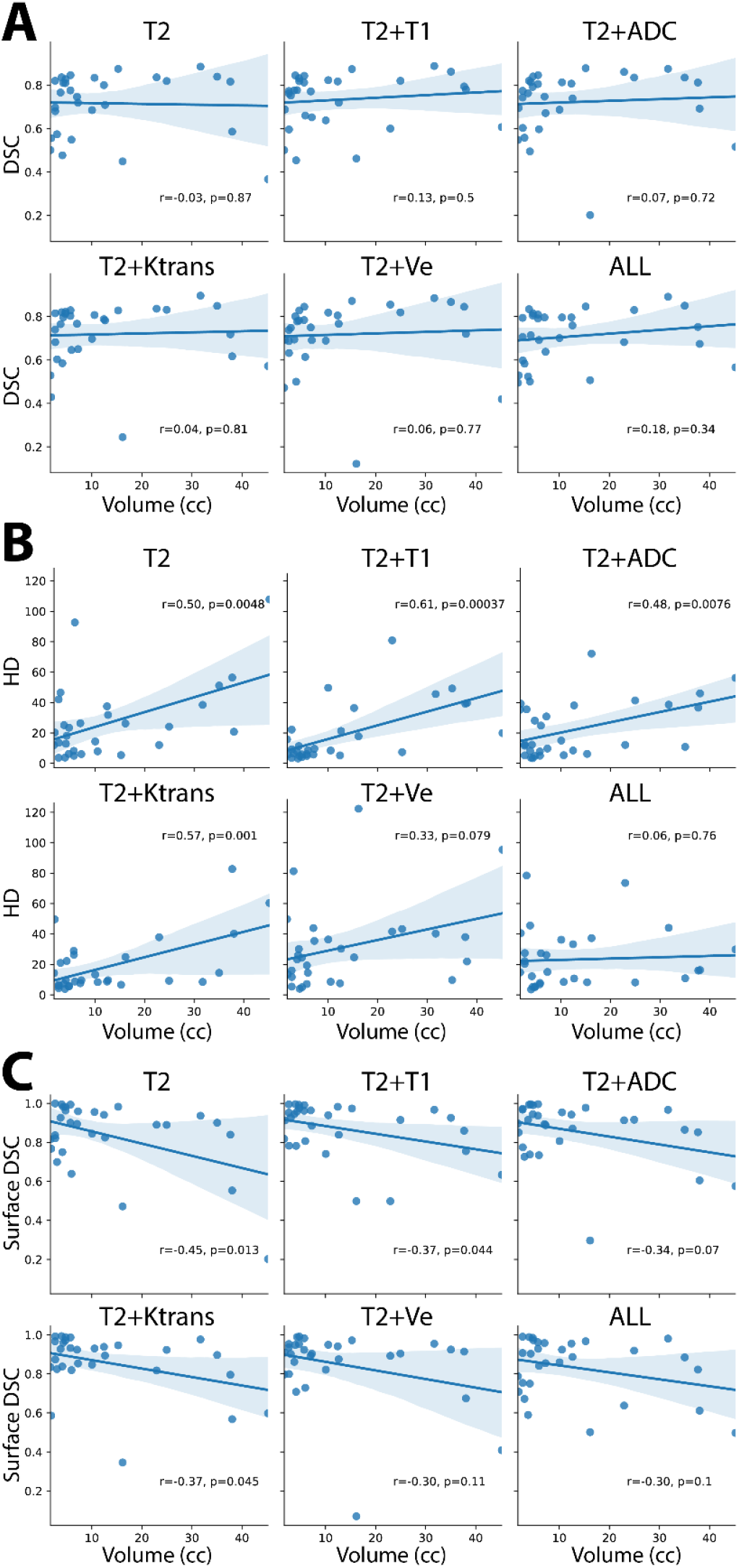
Dependence of tumor size on the Dice Similarity Coefficient (DSC) (**A**), Hausdorff Distance (HD) (**B**), and surface DSC (**C**), for various input channel models. T2=T2-weighted MRI, T1=T1-weighted MRI, ADC=apparent diffusion coefficient, Ktrans=volume transfer constant, Ve=extravascular extracellular volume fraction, ALL=all 5 input channels.

### 3.3. Clinical Evaluation

**Table 1** shows the categorical scores and predicted sources for ground truth and DL-generated (T2+T1 model) segmentations for each observer. The mean scores for ground truth vs. DL-generated segmentations were 3.0 vs. 2.5, 2.5 vs. 2.7, and 3.0 vs. 3.0 for observers 1, 2, and 3, respectively. Significance testing revealed no observer could differentiate between the scores (p>0.05) or source (p>0.05) of the ground truth segmentations compared to the DL-generated segmentations.

**Table 1.** Clinical evaluation and Turing test results for three physician expert observers. Each observer was asked to score blinded ground truth (GT) or deep learning (DL)-generated segmentations on a 4-point Likert scale (1 = requires corrections, large errors; 2 = requires corrections, minor errors; 3 = clinically acceptable, errors not clinically significant; 4 = clinically acceptable, highly accurate) and asked to identify the source of the segmentation (GT or DL). DL-generated segmentations corresponded to the best DL model tested (T2-weighted + T1-weighted).

| Observer | Score | GT (#) | DL (#) | p-value<sup>1</sup> | Source | GT (#) | DL (#) | p-value<sup>2</sup> |
| --- | --- | --- | --- | --- | --- | --- | --- | --- |
| **Observer 1**<br>**(Radiologist)** | 1 | 3 | 6 | 0.13 | GT | 16 | 10 | 0.18 |
|  | 2 | 7 | 11 |  | DL | 14 | 20 |  |
|  | 3 | 7 | 4 |  |  |  |  |  |
|  | 4 | 13 | 9 |  |  |  |  |  |
| **Observer 2**<br>**(Radiation-Oncologist)** | 1 | 3 | 6 | 0.44 | GT | 14 | 14 | 1.00 |
|  | 2 | 10 | 4 |  | DL | 16 | 16 |  |
|  | 3 | 16 | 13 |  |  |  |  |  |
|  | 4 | 1 | 7 |  |  |  |  |  |
| **Observer 3**<br>**(Radiation-Oncologist)** | 1 | 1 | 3 | 0.98 | GT | 9 | 12 | 0.61 |
|  | 2 | 8 | 6 |  | DL | 21 | 18 |  |
|  | 3 | 11 | 10 |  |  |  |  |  |
|  | 4 | 10 | 11 |  |  |  |  |  |

<sup>1</sup> Two-sided Wilcoxon signed rank tests were used for score comparisons. <sup>2</sup> McNemar tests were used for source prediction comparisons.

## 4. Discussion

In this pilot study, we determined the impact of mpMRI input channel combinations (T2, T2+T1, T2+ADC, T2+Ktrans, T2+Ve, ALL) on DL model segmentation performance. Recent work has suggested that the average agreement between physicians measured in DSC for OPC tumor segmentation is exceptionally low [44]. Notably, compared to previous fully-automated primary tumor segmentation studies of HNSCC patients, we achieved promising average DSC performance (**Table 2**). While it is difficult to directly compare DSCs between studies due to different datasets and model training, our models seemingly improve upon the only other fully-automated OPC tumor segmentation study to our knowledge (DSC=0.55), which exclusively investigated anatomical MRI [26].

**Table 2.**
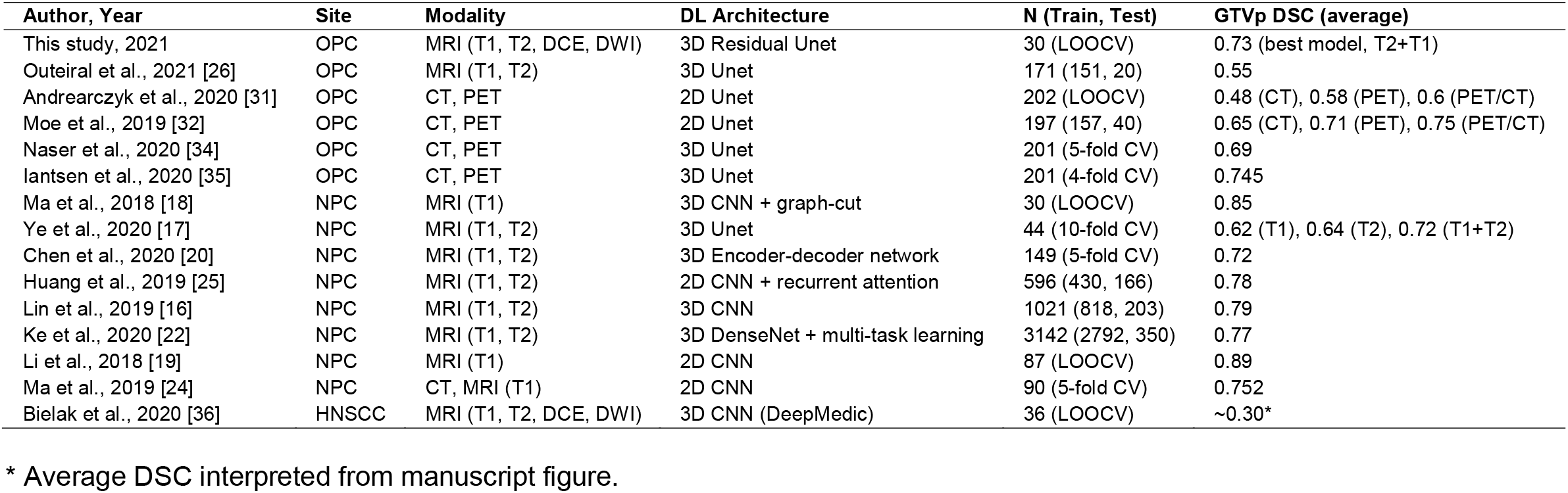
Survey of relevant DL auto-segmentation literature for comparison with our study. Only studies ≤3 years old and with sample sizes ≥30 were selected for comparison. DL=deep learning, N=number of images sets used in study, GTVp=primary gross tumor volume, DSC=Dice similarity coefficient, OPC=oropharyngeal cancer, NPC=nasopharyngeal cancer, HNSCC=head and neck squamous cell carcinoma, T2=T2-weighted MRI, T1=T1-weighted MRI, DCE=dynamic contrast enhanced MRI, DWI=diffusion weighted imaging MRI, LOOCV=leave-one-out cross-validation, CV=cross-validation, CNN=convolutional neural network.

The best average DSC performance was achieved by the T2+T1 model (DSC=0.73), which was higher than the baseline T2 model (DSC=0.72) but not statistically significant. Moreover, average DSC decreased when combining all input channels (DSC=0.71), though non-significantly. However, a previous similar study by Bielak et al. investigating HNSCC tumors with segmentations derived from T2 MRI demonstrated an increased DSC after the inclusion of all available mpMRI channels [36], which is in direct opposition to our results. Importantly, the authors used a smaller number of patients (n=18) than our study and implemented repeat imaging at different time-points, which could confound their results. Additionally, their results may be more relevant for a specific HNSCC tumor site, but no analysis was performed to verify this. Furthermore, it should be noted that the average DSC for their best model was ∼0.30, which was substantially lower than all our models. Notably, auto-segmentation studies in prostate cancer have also reported conflicting results on the additive effects of additional mpMRI input channels for DSC when using ground truth annotations derived from T2 MRI [47–49]. Therefore, further investigations are likely needed to verify if a significant positive DSC effect exists for mpMRI input channel combinations in OPC tumor auto-segmentation.

While most auto-segmentation studies have focused on DSC as an evaluation metric, it has been argued that other metrics should also be taken into consideration, depending on the use-case of the auto-segmentation tool [50,51]. Therefore, to increase the robustness of our analysis, we have included complimentary metrics (HD, FND, FPD, sensitivity, PPV, surface DSC, 95% HD, and MSD) to evaluate our models. Like DSC, most metrics show high performance across various models, with some models demonstrating significantly better values than the baseline T2 model. Interestingly, we demonstrated that in certain edge cases (low-performance example), the inclusion of additional channels could circumvent spurious voxel predictions derived from the baseline T2 model (a possible byproduct of model overfitting), which may increase model robustness. These results indicate that the additional channels may contain underlying additive information to improve performance for aspects other than traditional DSC-based evaluation. Notably, the specific anatomic subsite of the tumor (base of tongue or tonsil) had no significant effect on performance for any models for any evaluation metric, indicating that the models were robust to the spatial location of the OPC.

Previous studies [16,36] have suggested small tumors may be more difficult for DL models to segment, which would hinder the incorporation of models into radiotherapy workflows. Importantly, there were no significant correlations between tumor size and DSC for any of our models. However, it should be noted that surface distance metrics, such as the HD and surface DSC, demonstrate some size dependence, with larger and smaller tumors being easier for our models to segment, respectively. Interestingly, the surface distance metrics do not demonstrate a significant size dependence for some models that utilize additional channels, particularly those that correspond to functional parametric maps. Therefore, the inclusion of additional channels may strengthen the robustness of models to tumor size for surface distance metric performance, but further confirmatory work is needed.

The acceptability of segmentations used in a radiotherapy workflow is ultimately determined by physician judgment, with physician rating scales considered the gold standard for clinically relevant segmentation quality [51]. While subjective evaluation through rating scales is common in auto-segmentation studies, the established variability of OPC tumor segmentation between observers [44] highlights the difficulty in the interpretation of multi-observer segmentation quality analysis. Therefore, we implemented a comparative approach for each observer to determine if significant clinical differences were present between the ground truth segmentations and the corresponding segmentations of the best DL model (T2+T1). We demonstrated that experts were unable to determine differences between the ground truth and the DL-generated segmentations or identify the source of the segmentations. Therefore, our model “passed” the Turing test, which highlights its potential clinical utility. Of note, the radiologist who provided the original ground truth segmentations was the closest among the observers to correctly discriminating the segmentation sources but was still unable to achieve statistical significance. Moreover, for the radiation oncologist observers the mean clinical acceptability score of the DL-generated segmentations was equal to or higher than the ground truth segmentations, which may indicate a slight preference towards DL-generated OPC tumor segmentations for radiotherapy end users.

One limitation of our study is the use of a small cohort with standardized acquisition parameters. However, we have taken steps to optimally utilize our data by implementing a LOOCV approach and investigating various evaluation metrics. Moreover, we plan to include additional prospectively acquired data for model training and use external heterogenous validation sets in future studies to increase model generalizability. Another limitation of our study is that we have constrained our analysis of input image channels based on those that were investigated in previous literature [36]. However, mpMRI input channels can be further investigated through additional quantitative parametric maps (e.g., extended Tofts model [52], advanced DWI fitting models [53], etc.). Therefore, we plan to include additional input channels in future analyses. A final limitation of our study is the lack of overt image registration. Our images were acquired from a standardized clinical trial with patient immobilization; therefore, implicit co-registration was deemed adequate for tumor overlap. However, small amounts of motion artifacts may cause the segmentation mask to overlap improperly on mpMRI image channels, impacting auto-segmentation quality. Furthermore, though no geometric distortion was observed on any parametric maps, distortions were not explicitly quantified. Future studies should investigate the role of additional OPC-specific registration algorithms and geometric distortion correction in combination with mpMRI DL auto-segmentation algorithms.

## 5. Conclusions

In summary, using mpMRI inputs, we built OPC primary tumor DL auto-segmentation models that demonstrated excellent performance across multiple evaluation metrics, with average DSC scores as high as 0.73. Compared to our baseline model trained on T2 MRI only, we find that adding T1 MRI significantly improved HD, FND, sensitivity, surface DSC, and 95% HD. Moreover, adding Ve or using all input channels simultaneously significantly improved FND. Additionally, certain favorable aspects of model construction, including decreased spurious voxel predictions and robustness to tumor size when considering surface distance metric performance, are apparent for models that leverage additional input channels. Finally, physician experts could not differentiate ground truth from DL-generated segmentations, demonstrating our model “passed” the Turing test. These promising results should be further verified in large independent datasets. Overall, our pilot study is an important step towards fully automated MR-guided OPC radiotherapy workflows.

## Data Availability

Anonymized datasets are planned to be uploaded to a public repository in the near future. Until this time, anonymized data are available from the authors upon reasonable request.

## Acknowledgements

We thank Ms. Ann Sutton from the Editing Services Group at The University of Texas MD Anderson Cancer Center Research Medical Library for editing this article.

