## Supplementary Material for "Development of a High-Performance Multiparametric MRI Oropharyngeal Primary Tumor Auto-Segmentation Deep Learning Model and Investigation of Input Channel Effects: Results from a Prospective Imaging Registry"

**Supplementary Materials**

**Supplementary Methods**

Throughout our analysis, we adhered to the standardized artificial intelligence in medicine reporting guideline CLAIM [1] to ensure the reproducibility and rigor of our methodologic approach. The goal of this study was to create an oropharyngeal cancer (OPC) primary gross tumor volume (GTVp) auto-segmentation tool using multiparametric MRI (mpMRI) and determine the impact of individual mpMRI channels on model performance.

DICOM MRI images and DICOM RT GTVp structures were converted to Neuroimaging Informatics Technology Initiative (nifti) format for use in deep learning models using the DICOMRTTool v. 0.3.13 Python package [2].

*Model Architecture:* A DL convolutional neural network based on the 3D residual U-net architecture [3,4] was implemented in the Medical Open Network for Artificial Intelligence (MONAI) software package [5]. The images acted as variable-channel inputs to the models. The network consisted of four convolution blocks in the encoding and decoding branches and a bottleneck convolution block between the two branches. All convolution layers used a kernel size of 3 except for one convolution layer in the bottleneck, which used a kernel size of 1. The number of output channels for each convolution layer is shown in **Figure 1C** of the main text above each layer. Each convolution block in the encoding branch was composed of a two-strided convolution layer and a residual connection that contained a two-strided convolution layer and a one-strided convolution layer. In the bottleneck, the residual connection contained two one-strided convolution layers. In the decoding branch, each block contained a two-strided convolution transpose layer, a one-strided convolution layer, and a residual connection. Batch normalization and parametric rectified linear unit activation functions were used throughout the architecture, as parametric rectified linear unit functions have been found to improve upon the established rectified linear unit activation function, with increased performance for image-related tasks [6]. A softmax function was applied to the two-channel output to generate the GTVp segmentation mask (0 = background, 1 = tumor).

*Model Implementation:* Processed images were cropped to four random fixed-sized patches of size (96, 96, 96) per patch, with the center being considered foreground (i.e., GTVp, positive) or background (i.e., not GTVp, negative), with a 50% probability for both positive and negative cases. We implemented a batch size of two patients, resulting in a total of eight patches of images. The GTVp mask was used as the ground truth target to train the segmentation model. We implemented data augmentation on both the image and mask patches to mitigate overfitting, which included random horizontal flips of 50% and random affine transformations with an axial rotation range of 12 degrees and a scale range of 10%. Image processing and data augmentation were performed by the image transformation packages provided by the MONAI framework [5]. We implemented an Adam optimizer with a Sørensen-Dice similarity coefficient (DSC) loss function. The model was trained for 700 iterations with a learning rate of 2 x 10^-4^ for the first 550 iterations and 1 x 10^-4^ for the remaining 150 iterations.

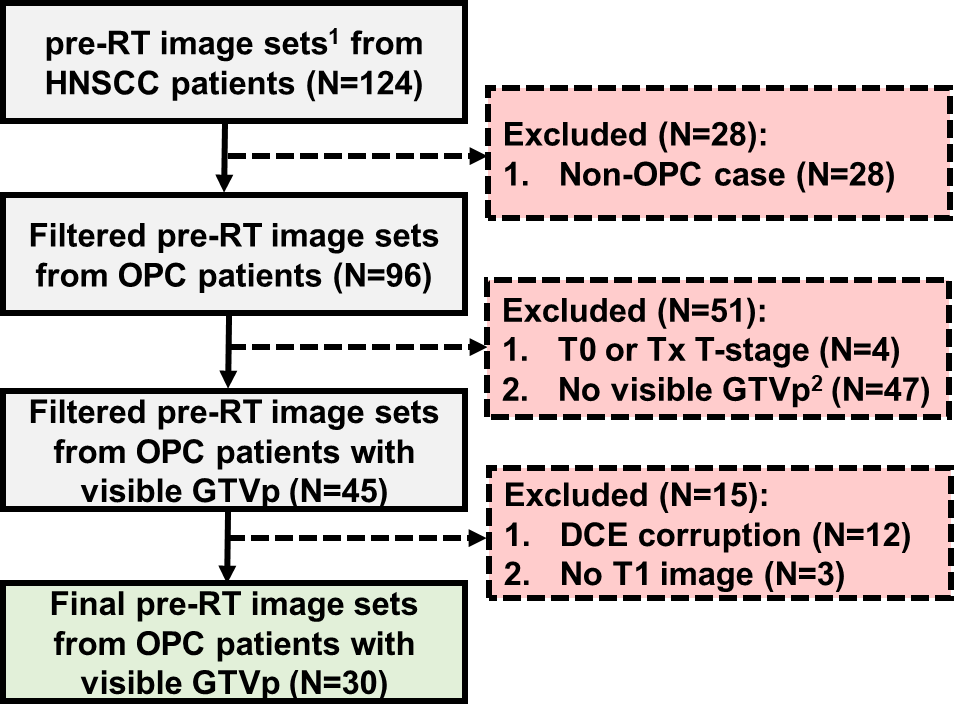

**Figure S1.** Patient selection flow diagram. ^1^ Images corresponded to T2-weighted, T1-weighted, dynamic contrasted enhanced, and diffusion weighted MRI sequences. ^2^ Tumors were not visible due to either surgery or induction chemotherapy before administration of radiotherapy. RT=radiotherapy, HNSCC=head and neck squamous cell carcinoma, OPC=oropharyngeal cancer, GTVp=primary gross tumor volume, DCE=dynamic contrast-enhanced MRI, T1=T1-weighted MRI, mpMRI=multiparametric MRI.

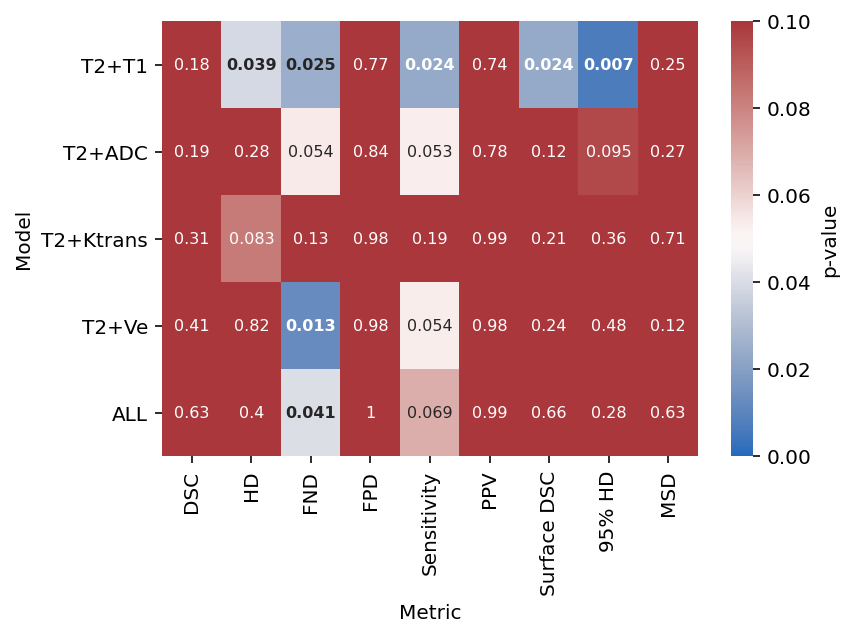

**Figure S2.** Heatmap of Wilcoxon signed-rank tests comparing additional input channel models with the baseline T2 model. Dice similarity coefficient (DSC), sensitivity, positive predictive value (PPV), and surface DSC comparison tests were one-way greater than, while Hausdorff distance (HD), false-negative DSC (FND), false-positive DSC (FPV), 95% HD, and mean surface distance (MSD) were one-way less than. Red corresponds to non-significant p-values, while blue corresponds to significant p-values. T2=T2-weighted MRI, T1=T1-weighted MRI, ADC=apparent diffusion coefficient, Ktrans=volume transfer constant, Ve=extravascular extracellular volume fraction, ALL=all 5 input channels.

**Table S1.** Patient demographic characteristics. Unless otherwise indicated, data shown correspond to patient number counts. HPV=human papillomavirus, AJCC=American Joint Committee on Cancer.

| **Characteristic** | **Value** |
| --- | --- |
| Age (median, range) | 65 (54-78) |
| Sex |  |
| Male | 30 |
| Female | 0 |
| Race |  |
| American Indian | 1 |
| Black/African American | 1 |
| White/Caucasian | 28 |
| Tumor subsite |  |
| Base of tongue | 10 |
| Tonsil | 20 |
| Tumor laterality |  |
| Right | 14 |
| Left | 12 |
| Bilateral | 3 |
| Midline | 1 |
| HPV status |  |
| Negative | 1 |
| Positive | 29 |
| T-category |  |
| T1 | 6 |
| T2 | 14 |
| T3 | 3 |
| T4 | 7 |
| N-category |  |
| N0 | 1 |
| N1 | 18 |
| N2a | 2 |
| N2b | 4 |
| N2c | 4 |
| N3 | 1 |
| AJCC stage |  |
| I | 10 |
| II | 5 |
| III | 6 |
| IVA | 8 |
| IVC | 1 |

**Table S2.** MRI sequence acquisition parameters. T2=T2-weighted MRI, T1=T1-weighted MRI, DCE=dynamic contrast enhanced MRI, DWI=diffusion weighted imaging MRI.

| **Acquisition Parameter** | **T2** | **T1** | **DCE** | **DWI** |
| --- | --- | --- | --- | --- |
| Repetition time (ms) | 4800.00 | 7.11 | 8.60 | 5400.00 |
| Echo time (ms) | 80.00 | 2.39 | 1.90 | 50.00 |
| Echo train length | 15 | 2 | 1 | 15 |
| Flip angle (°) | 180 | 10 | 15 | 120 |
| Slice thickness (mm) | 2.00 | 1.00 | 1.00 | 4.00 |
| In-plane resolution (mm) | 0.50 | 1.00 | 1.00 | 2.00 |
| Acquisition matrix | 256x230 | 256x256 | 128x83 | 128x128 |
| Pixel bandwidth (Hz/px) | 300.00 | 405.00 | 600.00 | 1220.00 |
| Number of averages | 1 | 2 | 1 | 8 |
| b-values (s/mm^2) | NA | NA | NA | 0, 800 |

**Table S3.** Mann-Whitney U tests between performance on base of tongue and tonsil subsites for each model for various evaluation metrics. All p-values were > 0.05. T2=T2-weighted MRI, T1=T1-weighted MRI, ADC=apparent diffusion coefficient, Ktrans=volume transfer constant, Ve=extravascular extracellular volume fraction, ALL=all 5 input channels.

| **Metric** | **Model** | **p-value** |
| --- | --- | --- |
| DSC | T2 | 0.404 |
| HD | T2 | 0.208 |
| FND | T2 | 0.404 |
| FPD | T2 | 0.214 |
| Sensitivity | T2 | 0.456 |
| PPV | T2 | 0.262 |
| Surface DSC | T2 | 0.404 |
| 95% HD | T2 | 0.346 |
| MSD | T2 | 0.456 |
| DSC | T2+T1 | 0.491 |
| HD | T2+T1 | 0.387 |
| FND | T2+T1 | 0.184 |
| FPD | T2+T1 | 0.371 |
| Sensitivity | T2+T1 | 0.291 |
| PPV | T2+T1 | 0.346 |
| Surface DSC | T2+T1 | 0.208 |
| 95% HD | T2+T1 | 0.387 |
| MSD | T2+T1 | 0.422 |
| DSC | T2+ADC | 0.491 |
| HD | T2+ADC | 0.354 |
| FND | T2+ADC | 0.284 |
| FPD | T2+ADC | 0.184 |
| Sensitivity | T2+ADC | 0.371 |
| PPV | T2+ADC | 0.241 |
| Surface DSC | T2+ADC | 0.172 |
| 95% HD | T2+ADC | 0.202 |
| MSD | T2+ADC | 0.291 |
| DSC | T2+Ktrans | 0.491 |
| HD | T2+Ktrans | 0.430 |
| FND | T2+Ktrans | 0.338 |
| FPD | T2+Ktrans | 0.189 |
| Sensitivity | T2+Ktrans | 0.354 |
| PPV | T2+Ktrans | 0.422 |
| Surface DSC | T2+Ktrans | 0.208 |
| 95% HD | T2+Ktrans | 0.131 |
| MSD | T2+Ktrans | 0.422 |
| DSC | T2+Ve | 0.474 |
| HD | T2+Ve | 0.322 |
| FND | T2+Ve | 0.330 |
| FPD | T2+Ve | 0.338 |
| Sensitivity | T2+Ve | 0.422 |
| PPV | T2+Ve | 0.371 |
| Surface DSC | T2+Ve | 0.387 |
| 95% HD | T2+Ve | 0.322 |
| MSD | T2+Ve | 0.474 |
| DSC | ALL | 0.404 |
| HD | ALL | 0.122 |
| FND | ALL | 0.322 |
| FPD | ALL | 0.255 |
| Sensitivity | ALL | 0.422 |
| PPV | ALL | 0.262 |
| Surface DSC | ALL | 0.422 |
| 95% HD | ALL | 0.430 |
| MSD | ALL | 0.354 |
